# Air pollution and out-of-hospital cardiac arrest risk

**DOI:** 10.1101/2023.03.15.23287335

**Authors:** L. Moderato, D. Aschieri, D. Lazzeroni, L. Rossi, S. Bricoli, A. Biagi, S. Ferraro, S.M. Binno, A. Monello, V. Pelizzoni, C. Sticozzi, A. Zanni, G. Magnani, F.L. Gurgoglione, A. Capucci, S. Nani, R.A. Montone, D. Ardissino, F. Nicolini, G. Niccoli

**Affiliations:** Cardiology Department, “Guglielmo da Saliceto” Hospital, Piacenza, Italy; IRCCS Fondazione Don Carlo Gnocchi, Firenze, Italy; Cardiology Unit, Department of Medicine and Surgery, University of Parma, Italy; Abcardio, Bologna, Italy; Emergency Department, “Guglielmo da Saliceto” Hospital, Piacenza, Italy; Department of Cardiovascular Sciences. Fondazione Policlinico Universitario A. Gemelli IRCCS, Rome, Italy; Cardiac Surgery Unit, Department of Medicine and Surgery, University of Parma, Italy

## Abstract

**Background:** Globally nearly 20% of cardiovascular disease deaths were attributable to air pollution. Out-of-hospital cardiac arrest (OHCA) represents a major public health problem, therefore, the identification of novel OHCA triggers is of crucial relevance. The aim of the study was to evaluate the association between air pollution (short-, mid-and long-term exposure) and out-of-hospital cardiac arrest (OHCA) risk, during a 7 years-period from a highly polluted urban area with a high density of automated external defibrillators (AEDs).

**Methods and results:** OHCA were prospectively collected from the “Progetto Vita Database” between 01/01/2010 to 31/12/2017; day-by-day air pollution levels were extracted from the Environmental Protection Agency (ARPA) stations. Electrocardiograms of OHCA interventions were collected from the AEDs data cards. Day-by-day particulate matter (PM) 2.5 and 10, ozone (O3), carbon monoxide (CO) and nitrogen dioxide (NO2) levels were measured. A total of 880 OHCAs occurred in 748 days. A significantly increased in OHCA risk with the progressive increase in PM 2.5, PM 10, CO and NO2 levels was found. After adjustment for temperature and seasons, a 9% and 12% increase of OHCA risk for each 10 μg/m3 increase of PM 10 (p< 0.0001) and PM 2.5 (p< 0.0001) levels was found. Air pollutants levels were associated with both asystole and shockable rhythm risk while no correlation was found with pulseless electrical activity.

**Conclusions:** Short-term and mid-term exposure to PM 2.5 and PM 10 is independently associated with the risk of OHCA due to asystole or shockable rhythm.

## INTRODUCTION

For decades air pollution and its potential pathogenetic effects on people’s health have been underestimated both by governments and physicians, but nowadays, air pollution represents the largest environmental cause of disease and death worldwide^1^. The World Health Organization estimates that around 7 million people die every year from exposure to polluted air since 91% of the world’s population lives in areas where air contaminants exceed safety levels^2^. For these reasons, in a recent joint opinion paper, the World Heart Federation, American College of Cardiology, American Heart Association, and the European Society of Cardiology have stated that globally nearly 20% of cardiovascular disease deaths were attributable to air pollution^2,3^. Furthermore, air pollution was the 4th highest-ranking risk factor for mortality, with more attributable deaths than high LDL cholesterol, high body-mass index, physical inactivity or alcohol use^2^.

Air pollution is a complex and dynamic mixture of numerous compounds in gaseous and particle form, originating from diverse sources, subject to atmospheric transformation and varying over space and time. The common air pollutants monitored by public security systems are particulate matter 2.5 (PM 2.5) and 10 (PM 10), ozone (O_3_), carbon monoxide (CO) and nitrogen dioxide (NO_2_). PM is the most thoroughly studied component of air pollution and has been strongly associated with multiple diseases and pathogenic mechanisms^4^.

Several potential pathophysiological mechanisms linking air pollution to cardiovascular disease and death have been demonstrated, including endothelial dysfunction, autonomic dysfunction, inflammation, hypertension and hypercoagulability, that all together may potentially lead to plaque instability, coronary thrombosis and eventually life-threatening ventricular arrhythmias ^3,5,6^. Both short-term (from hours to days) and long-term (from months to years) exposure to ambient air pollutants has been associated with cardiovascular disease and death^7^. Short-term variations in PM 2.5 levels have been associated with increased myocardial infarction, stroke, and cardiovascular death risk^2,4,8^.

Out-of-hospital cardiac arrest (OHCA) represents a dramatic event and a major public health challenge. OHCA incidence in Europe is estimated at 275.000/year^9^. Despite the recent efforts to improve resuscitation, such as early defibrillation and fast intervention, OHCA survival remains low (ranging from 9% to 20% at hospital discharge and up to 50% if the return of spontaneous circulation is achieved)^10–12^. Therefore, the identification of novel OHCA triggers is of crucial relevance in preventing such a dramatic event. Recently, intermediate (21 days) PM exposure has been associated with ventricular arrhythmias in high selected population such as high-risk patients with dual-chamber implanted cardioverter defibrillator^13^. However, the relation between OHCA and air pollutants is still an open issue, in particular in Europe, as many previous studies derived from China or the Eastern world, with a different epidemiological and environmental background. The present study aimed to evaluate the influence of both short, mid and long-term exposure to air pollutants on OHCA risk in a small and highly polluted Italian urban area.

## METHODS

### “Progetto Vita Registry”

“Progetto Vita” is an independent, donations and citizen volunteers supported no-profit association based in Piacenza that supplements standard, government-funded emergency medical system (EMS) when responding to OHCA. The “Progetto Vita” project was initiated on June 6 1999, shortly after automated external defibrillators (AEDs) became available in Europe as a low-cost, simplified option for quickly training laypersons without the need for traditional, more expensive and time-consuming basic life support and defibrillation courses. The original structure and organization of “Progetto Vita” have been previously reported (www.Progetto-vita.eu)^14^. Over the next 20 years, “Progetto Vita” grew organically through the dedicated effort to recruit more than 25,000 citizen volunteers. Phone calls to EMS (#118 or #112 in EU, equivalent to #911 in the United States) simultaneously activate “Progetto Vita” and the standard EMS system. Over time, “Progetto Vita” progressively increased the availability of AEDs in the public places of Piacenza from their initial numbers of 21 in 1999 to 618 by December 31, 2017 (in public venues and police or fire-fighter vehicles), thereby allowing a high AEDs density into the urban area (a mean of 5 defibrillators for every km^2^). Efficacy of “Progetto Vita” in both promoting early defibrillation and improving OHCA survival have been previously reported.^14^

### sOut-of-hospital Cardiac arrest definition

All OHCA cases of residents in Piacenza City between 2010 and 2017 were prospectively collected and included in the “Progetto Vita Registry”. OHCA was defined as the sudden cessation of cardiac mechanical activity in the absence of a non-cardiac cause of cardiac arrest. OHCA definition was checked by the Authors and based on clinical judgement, EMS and bystander information and medical registries. Traumatic death, as well as terminally ill cardiac arrests, were excluded. Syncope and other forms of transient loss of consciousness were also excluded. Data regarding OHCA patients’ gender, age, location of cardiac arrest, and time were extracted from Progetto Vita and EMS registry. Electrocardiograms of OHCA interventions were collected from the data cards housed in all the AEDs. Electrocardiograms were classified as asystole, pulseless electrical activity, or shockable rhythm (ventricular tachycardia or ventricular fibrillation)^14^; asystole and/or shockable rhythm were considered as a cause of OHCA, while pulseless electrical activity was considered as a primarily non-cardiac cause of arrest.

### Air pollution and meteorological data

Piacenza covers a territory of 118.5 km^2^ with a population of 102.355 inhabitants. Exposure data were provided by the Regional Environmental Protection Agency (ARPA) local monitoring stations (code 05000033 and 05000065). Air pollution levels were collected independently and blindly of outcome data and then merged. All stations hourly monitored PM 2.5, PM 10, NO2, CO and O3 levels. The daily mean concentrations of the pollutants were calculated and no data were missing. 7-days, 21-days and 1-year pollutant exposure were calculated as the mean concentration of each pollutant (mean exposure) during the previous 7, 21 and 365 days preceding the OHCA event. Meteorological data including hourly values of temperature were obtained from the urban meteorological stations. Only OHCAs within the urban area (10 km range from the monitoring station) were selected. This study was approved by the regional ethical review board and complied with the Declaration of Helsinki.

### Statistical analysis

Continuous variables were expressed as mean and standard deviation and differences among groups were tested by ANOVA, with Least-Significant Difference post-hoc analysis. Categorical variables were expressed as a number and percentages testing differences among groups by the Pearson chi-square test. A time-stratified case-crossover design selection coupled with conditional logistic regression analysis was used in order to assess independent predictors of OHCA. The time-stratified case-crossover design was first proposed in 1991 by Maclure^15^ and is a well-established study design to assess the association between transient exposure to air pollution and emergency health events; it is a common design in air pollution epidemiological studies^4,16,17^ that was used to analyse air pollution and the risk of OHCA. The case day was defined as the day of the OHCA occurrence, while the days without event were considered as controls. Short-term (24 hours), mid-term (7 days and 21 days) and long-term (365 days) mean exposure prior to a given event (case) was compared with the mean exposure during the corresponding periods without events (controls). The estimate risk was expressed as Odds ratio (OR) with a 95% confidence interval (CI). The crude models were extended with multiple adjustments for temperature and seasons. Statistical significance was set at p <0.05. All statistics were performed with SPSS version 24 (IBM Corporation, Armonk, NY).

## Results

A total of 880 OHCA cases from January 2010 to December 2017 occurred during 748 OHCA days. Days with 1 OHCA were 748 (26%), days with 2 OHCA were 102 (3.5%) and days with 3 OHCA were 15 (0.5%). Ventricular fibrillation was recorded in 141 patients (16%) while 739 (84%) OHCAs showed non-shockable rhythm (20 pulseless electrical activities and 719 cases of asystole). The overall survival rate was 2.2%, while the survival rate of OHCAs with shockable rhythm was 33%. Mean age was 76 ± 15 and 55% of patients were male (484).

Air pollutants mean concentrations (overall and divided according to different seasons) are shown in Table 1. PM 2.5 and PM 10 exceeded safety limits (> 25 μg/m3 and >50 μg/m3, respectively) for 443 (15.9%) and 482 (16.5%) days. CO mean concentration was 5.3±3.1 mg/m^3^ and during 282 days (9.7%) CO level exceeded safety limit (>10 mg/m3). NO_2_ mean concentration was 42±20 µg/m3, with 1292 days (45%) over safety limit. Finally, O_3_ mean concentration was 43±33 μg/m^3^, with only 5.5% days over the safety limit (>100 μg/m^3^).

**Table 1.** Air pollutants concentrations according to different seasons

|  | Overall | Summer | Autumn | Winter | Spring | p value |
| --- | --- | --- | --- | --- | --- | --- |
| **PM 2.5 (mean ± DS)** | 23 ± 16 mcg/m<sup>3</sup> | 15±5mcg/m<sup>3</sup> | 15± 7 mcg/m<sup>3</sup> | 35±19mcg/m<sup>3</sup> | 29±19mcg/m<sup>3</sup> | <0.001 |
| **PM 10 (mean ± DS)** | 33 ± 20 mcg/m<sup>3</sup> | 22 ±8 mcg/m<sup>3</sup> | 25 ± 11 mcg/m<sup>3</sup> | 46 ± 24 mcg/m<sup>3</sup> | 42 ± 21 mcg/m<sup>3</sup> | <0.001 |
| **CO (mean ± DS)** | 5.32 ± 3.14 mcg/m<sup>3</sup> | 2.81 ± 0.09 mcg/m<sup>3</sup> | 3.84 ± 1.35 mcg/m<sup>3</sup> | 8.43 ± 2.85 mcg/m<sup>3</sup> | 6.35 ± 3.10 mcg/m<sup>3</sup> | <0.001 |
| **NO<sub>2</sub> (mean ± DS)** | 42 ± 20 mcg/m<sup>3</sup> | 24 ± 7 mcg/m<sup>3</sup> | 35 ± 11 mcg/m<sup>3</sup> | 63 ± 21 mcg/m<sup>3</sup> | 47 ± 14 mcg/m<sup>3</sup> | <0.001 |
| **O<sub>3</sub> (mean ± DS)** | 43 ±33 mcg/m<sup>3</sup> | 79 ± 23 mcg/m<sup>3</sup> | 61 ± 19 mcg/m<sup>3</sup> | 16 ± 14 mcg/m<sup>3</sup> | 16 ± 14 mcg/m<sup>3</sup> | <0.001 |

Legend. OHCA, out-of-hospital cardiac arrest; PM, particulate matter; O<sub>3</sub>, ozone; CO, carbon monoxide; NO<sub>2</sub>, nitrogen dioxide.

### Short-term exposure to air pollution and out-of-hospital cardiac arrest risk

Concentrations of PM 2.5, PM 10, CO and NO_2_ were significantly higher in OHCA days compared to non-OHCA days (Table 2). A significant increased risk of OHCA with the progressive increase in PM 2.5, PM 10, CO and NO_2_ levels was found; all correlations between OHCA risk and air pollutant levels are shown in Figure 1 and Table S1. More specifically, OHCA risk significantly increased with the progressive increase of PM 2.5, corresponding to a 12% increase of the relative risk of OHCA for each 10 μg/m3 increase of PM 2.5 levels (OR=1.012; 1.007-1.017; p<0.0001) (figure 2). Similarly, OHCA risk significantly increased with the progressive increase of PM 10 corresponding to a 9% increase of the relative risk of OHCA for each 10 μg/m3 increase of PM 10 levels (OR=1.009; 1.005-1.013; p< 0.0001) (figure 3).

**Table 2.**
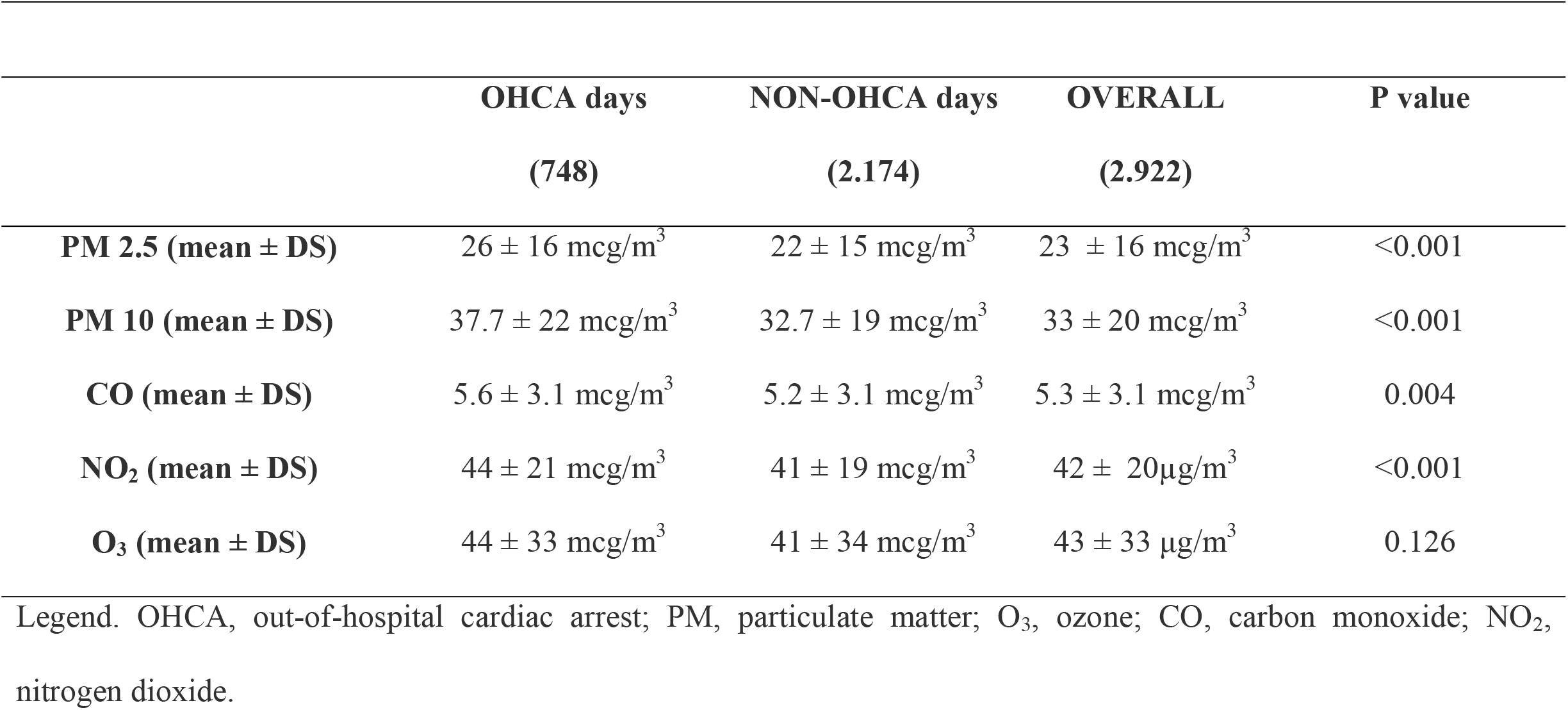
Air pollutions levels in days with or without OHCA

**Figure 1.**
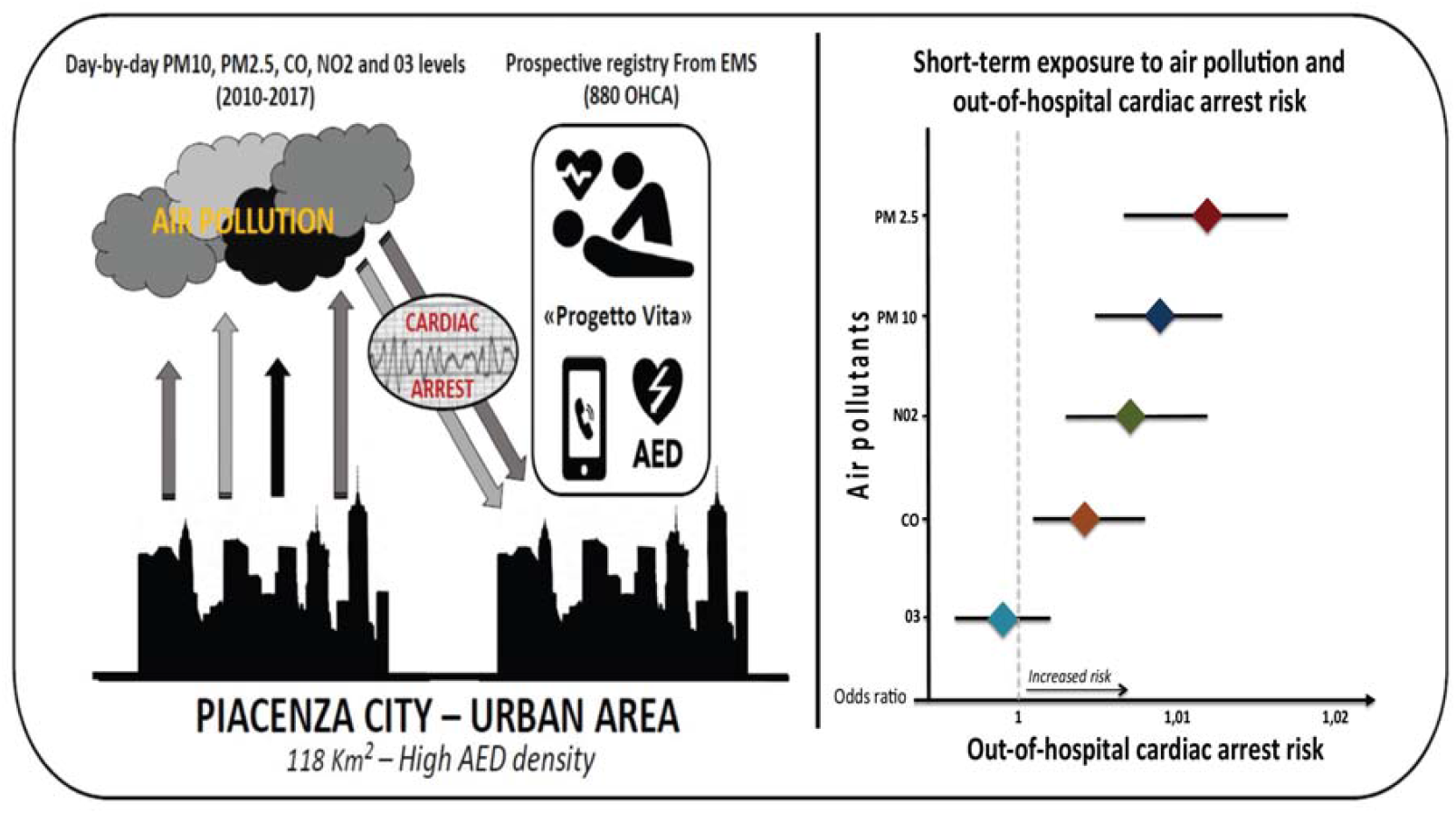
(Central figure). The study investigated the association between day-by-day urban PM10, PM2.5, CO, NO2 and O3 levels and the risk of out-of-hospital cardiac arrest (OHCA) during a 7 years-period in a large cohort of patients from a highly polluted urban area with a high density of automated external defibrillators (AEDs). A total of 880 OHCAs were prospectively collected from the “Progetto Vita Database” of cardiac arrest between 01/01/2010 to 31/12/2017 (left side). A significantly increased risk of OHCA with the progressive increase in PM 2.5, PM 10, CO and NO2 levels was found (right side).

**Figure 2.**
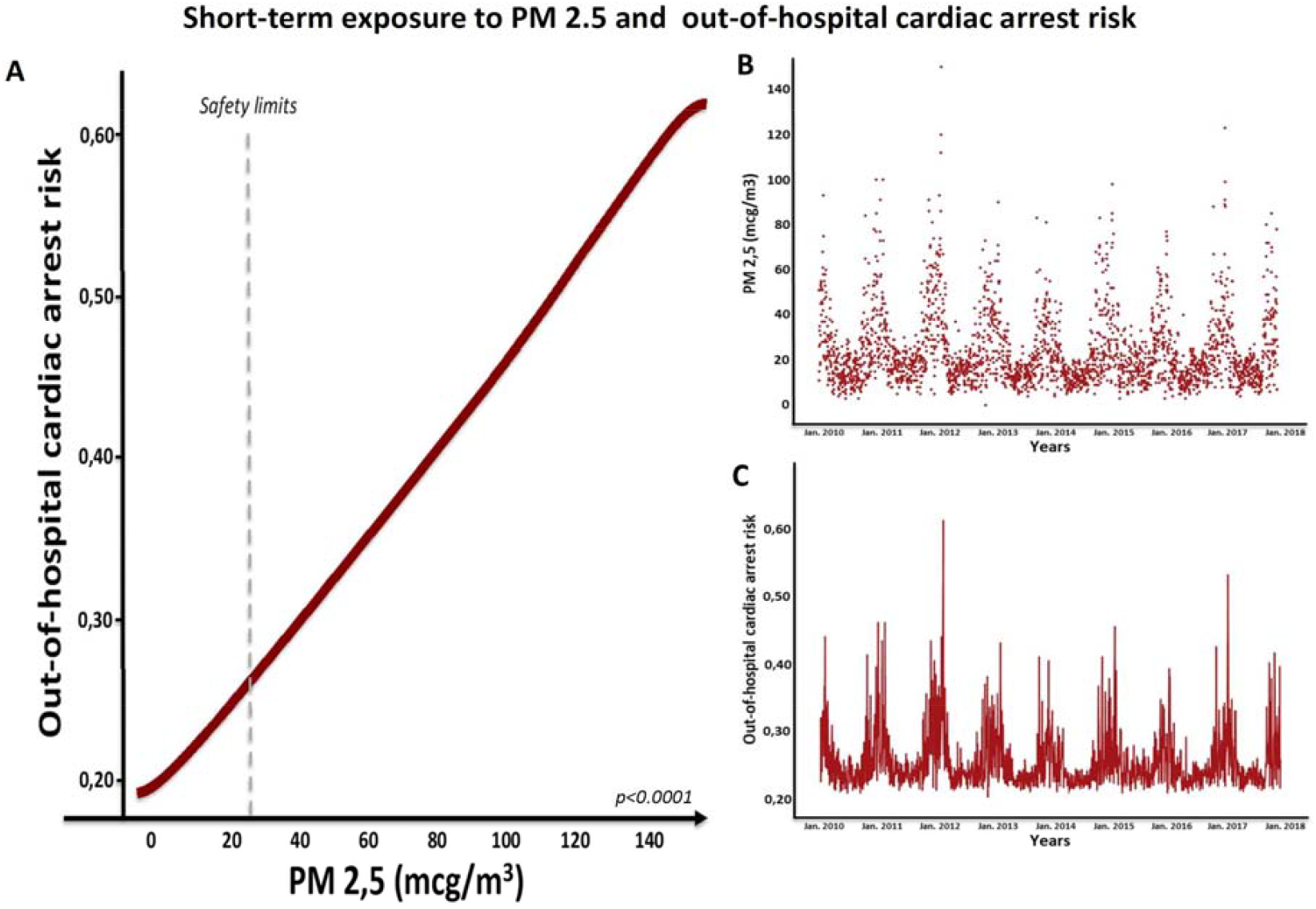
Short-term (same day) PM 2.5 levels and out-of-hospital cardiac arrest (OHCA) risk (panel A). Daily PM 2.5 levels from 01/01/2010 to 31/12/2017 (panel B); panel C shows the day-by-day distribution of the OHCA risk associated with each corresponding daily PM 2.5 value, from from 01/01/2010 to 31/12/2017.

**Figure 3.**
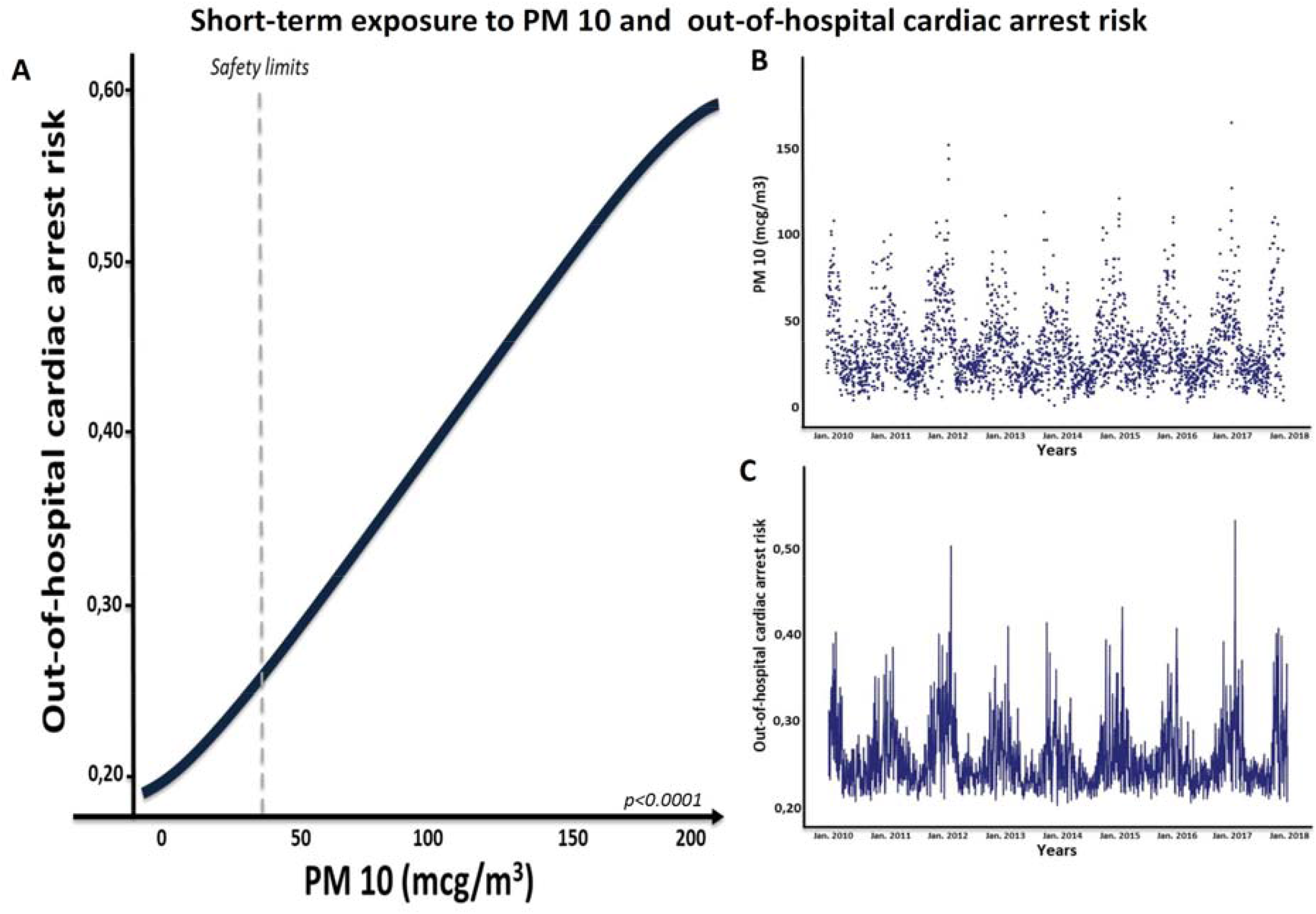
Short-term (same day) PM 10 levels and out-of-hospital cardiac arrest (OHCA) risk (panel A). Daily PM 10 levels from 01/01/2010 to 31/12/2017 (panel B); panel C shows the day-by-day distribution of the OHCA risk associated with each corresponding daily PM 10 value, from from 01/01/2010 to 31/12/2017.

Moreover, OHCA risk significantly increased with the progressive increase of CO, corresponding to a 4.5% increase of the relative risk of OHCA for each mg/m^3^ increase of CO levels (OR=1.456; 1.124-1.886; p<0.04). OHCA risk significantly increased with the progressive increase of NO_2_, corresponding to a 7% increase of the relative risk of OHCA for each 10 μg/m^3^ increase of NO_2_ levels (OR=1.007; 1.003-1.0012; p<0.001). Finally, OHCA risk significantly increased with the progressive increase of NO_2_, corresponding to a 7% increase of the relative risk of OHCA for each 10 μg/m^3^ increase of NO_2_ levels (OR=1.007; 1.003-1.0012; p<0.001). No significant increased risk of OHCA was found considering the progressive increase of O_3_ levels (OR=0.999; 0.996-1.002; p=0.577).

Interestingly, higher mean air pollutants (PM 2.5, PM 10, CO and NO_2_) daily values were found with the progressive increase of OHCA number per day (Figure 4). The correlation between PM 2.5, PM 10, CO and NO_2_ levels OHCA risk remained significant even considering 7 days before the OHCA event (Table S1 and Figure S1). Moreover, the correlation between PM 2.5, PM 10 and NO_2_ levels and OHCA risk remained significant even considering separately both male and female as well as age above or below 65 years (Table S2).

**Figure 4.**
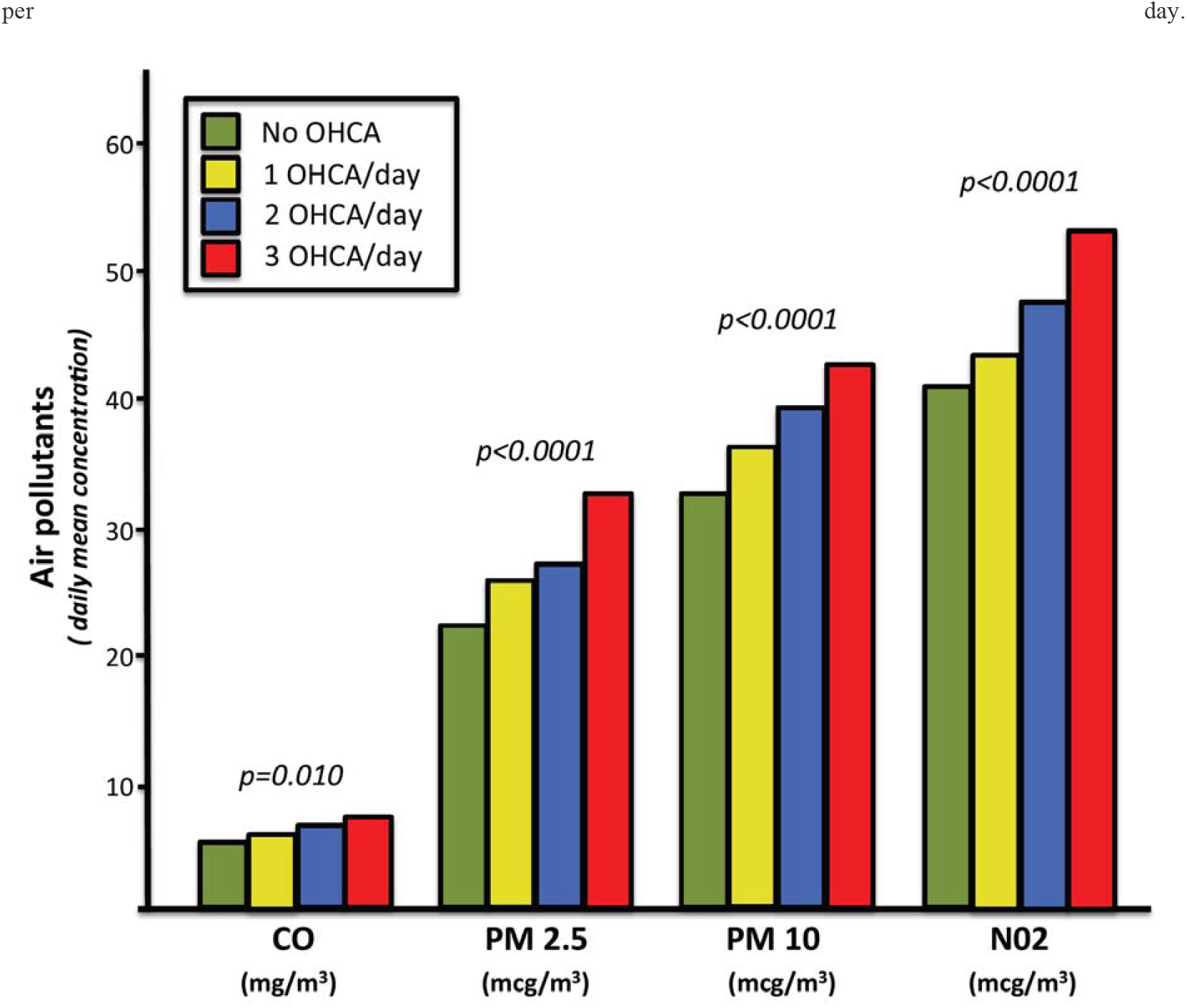
Increase in the average air pollution levels associated with the progressive increase in the number of OHCAs per day.

After adjustment for temperature and seasons, only the correlation between PM 2.5 and PM 10 levels and OHCA risk remained significant (OR=1.012; 1.006-1.021; p<0.0001 and OR=1.010; 1.004-1.016; p<0.001 and, respectively), while the correlation between CO and NO_2_ levels failed to remain independently associated.

### Mid-and Long-term exposure to air pollution: out-of-hospital cardiac arrest risk

The correlation between PM 2.5, PM 10, CO and NO_2_ levels and OHCA risk remained significant even considering 7-days and 21-days intervals before the OHCA event (Figure S1 - table S1). Conversely, no correlations between air pollutants levels and OHCA risk was found considering a longer period of exposure (365-days before the OHCA event - Figure S1, Table S2).

### Air pollutant safety limits and out-of-hospital cardiac arrest risk

A higher OHCA rate was found in days with PM 2.5 over the safety limit (23% vs 29%; p=0.001), corresponding to a 1.38-fold higher risk of OHCA (OR 1.38; 1.11-1.73; p=0.001). Similarly, a higher OHCA rate was found in days with PM 10 over safety limit (24% vs 31%; p=0.005), corresponding to a 1.36-fold higher risk of OHCA (OR 1.36; 1.01-1.68; p=0.005) (Figure 5). Moreover, a higher OHCA rate was found in days with NO_2_ above the safety limit (50% vs 43%. p=0.002), corresponding to a 1.31-fold higher risk of OHCA (OR 1.31; 1.11-1.55; p=0.002). A trend towards a significantly higher OHCA rate was found in days with CO above the safety limit (12% vs 9%. p=0.061) and no differences in OHCA’s rate were found considering days with O_3_ levels above safety limits (6.1 vs 5.3%. p=0.449).

**Figure 5.**
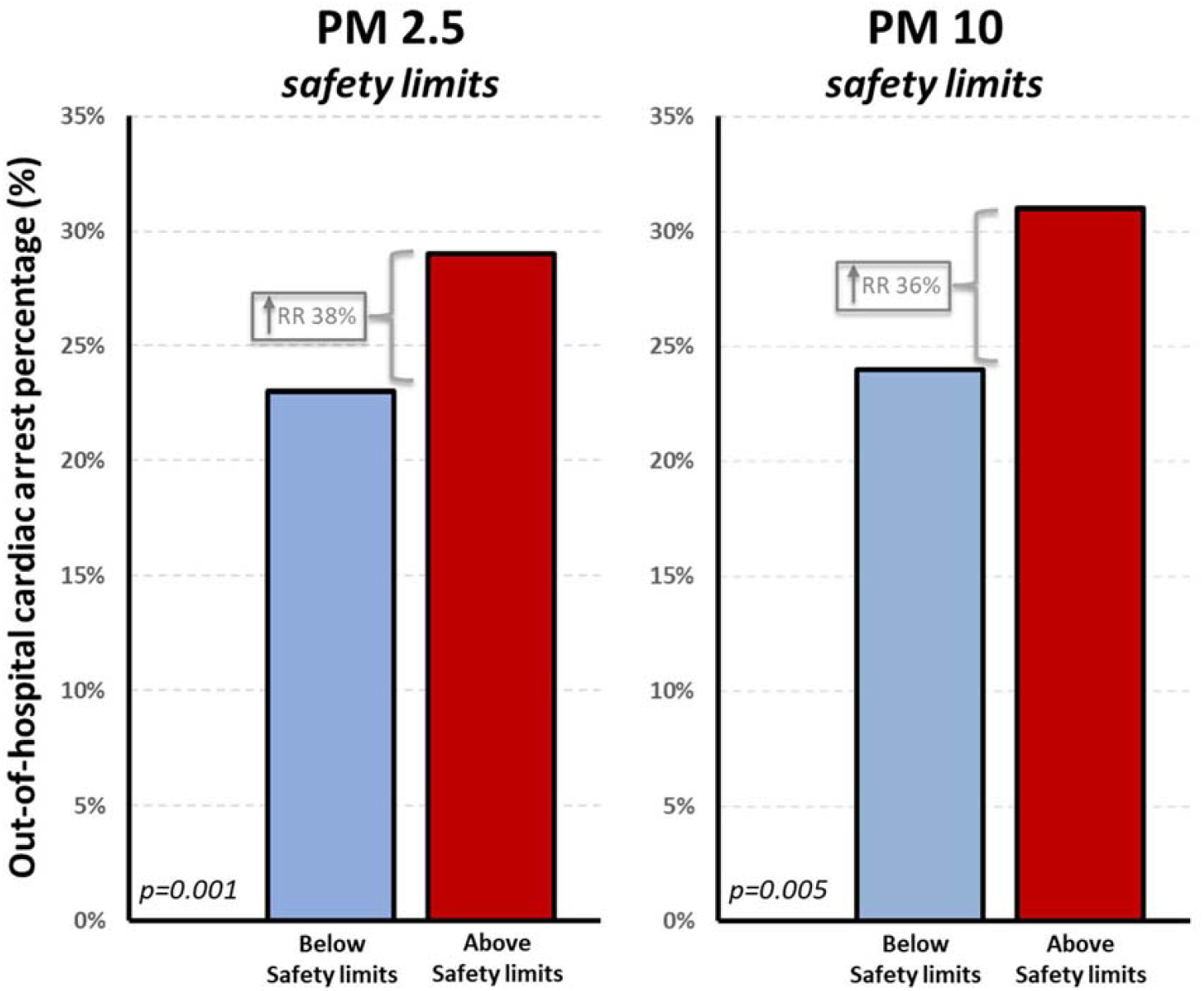
Higher rate of OHCA during the days in which the air pollutants (PM 2.5 and PM 10) exceed WHO safety levels with associated corresponding increase in the relative risk of OHCA.

### Electrocardiograms of OHCA interventions and out-of-hospital cardiac arrest risk

PM 2.5, PM 10, NO_2_ and CO levels were associated with an increased risk of asystole or ventricular fibrillation (Figure 6). Of note, pulseless electrical activity risk (a rhythm associated with non-primary cardiac cause of out-of-hospital arrest) was not correlated to any air pollutant levels.

**Figure 6.**
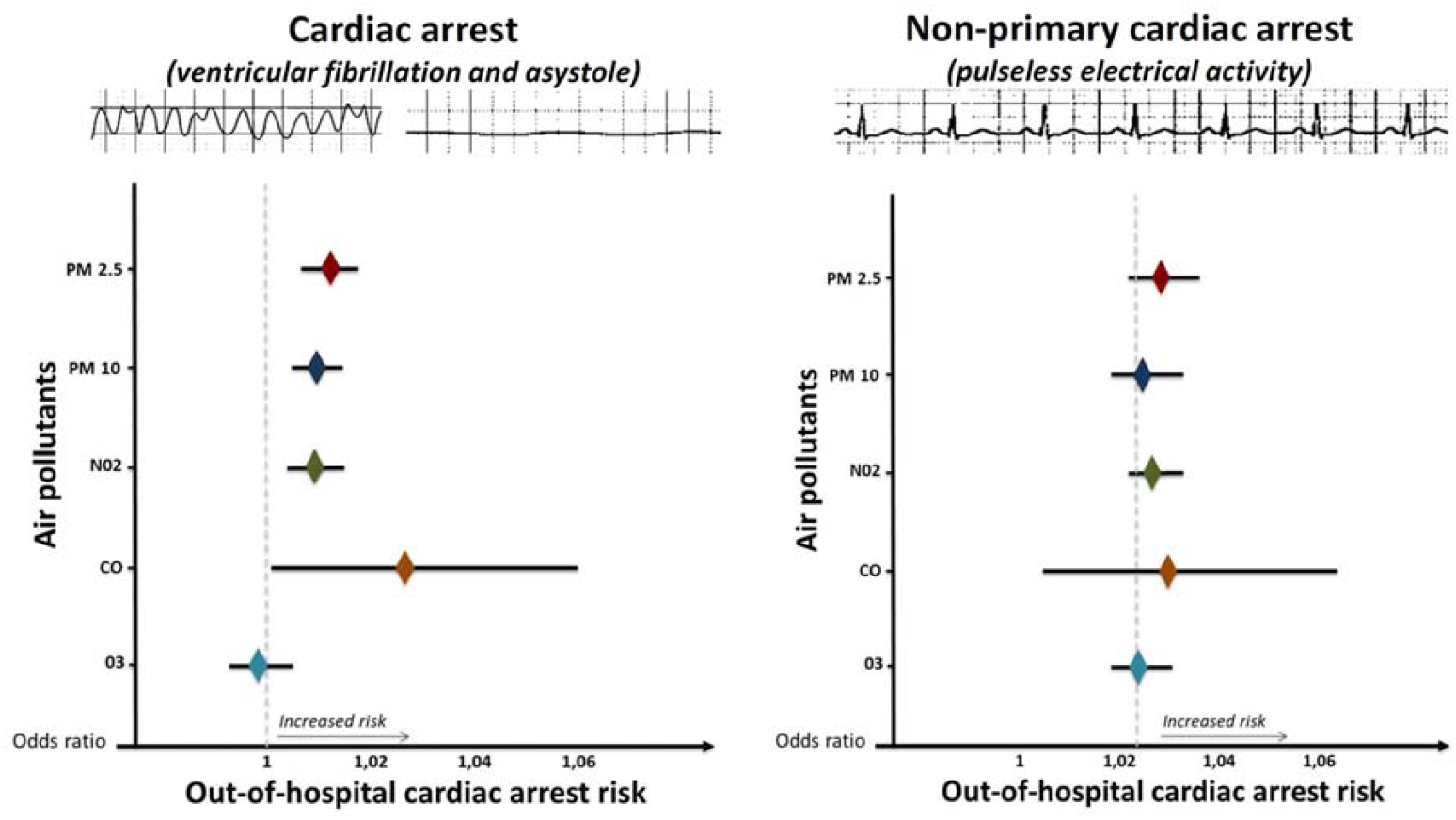
Air pollutants and out-of-hospital cardiac arrest (OHCA) risk according to different AEDs rhythm: ventricular fibrillation and asystole vs pulseless electrical activity (non-primary cardiac arrest).

## Discussion

The present study demonstrates a significant association between OHCA risk and short- and mid-term exposure to PM 2.5, PM 10. In particular, a 12% increased relative risk of OHCA for each 10 μg/m^3^ PM 2.5 levels as well as a 9% increased relative risk of OHCA for each 10 μg/m^3^ increase of PM 10 levels were found. Of note, PM 2.5 and PM 10 levels only remained significantly correlated to OHCA risk, after adjustment for temperature and seasons. Interestingly, in days with PM exceeded safe limits, an increase in relative risk of OHCA of 38% was found considering PM 2.5 levels as well as an increase in relative risk of OHCA of 36% was found considering PM 10. On the other hand, no correlation was found between all air pollutants and AEDs detected rhythm associated with primarily non-cardiac cause of OHCA, such as pulseless electrical activity, thus suggesting that air pollutants levels are associated with increased risk of cardiac causes of OHCA only.

Since 2015, the expert position paper on air pollution and cardiovascular disease reported a direct link between air pollutants and cardiovascular diseases and death^3^. The correlation between air pollution and cardiovascular diseases was found to be most consistent considering PM, rather than CO, NO_2_ and O_3_, which is responsible for the vast majority of the disease burden via its impacts on ischemic heart disease^3,5,18^and stroke^19^. Our results, showing a stronger and independent correlation between OHCA and PM, are consistent with previous reports, even considering OHCA as a cause of cardiac death.

In particular, time-series studies conducted in hundreds of urban areas globally showed a consistent association between short-term variability in PM and cardiovascular disease deaths, while large cohort studies from both high and lower-income settings demonstrated an increased cardiovascular disease incidence and mortality in association with PM levels^4^. Multiple links between PM and cardiovascular diseases have been proposed: alterations in vascular tone and increased blood pressure^18^, coronary artery calcification, plaque instability^20^, left ventricle hypertrophy and type 2 diabetes^21^. Furthermore, PM air pollution has been associated with the progression of atherosclerosis^22^.

A recent review^23^ highlighted that the risk of cardiovascular events increases by 0.1 to 1.0% for each short-term increment of a cubic millimetre in the PM 2.5 level; our results were similar since a 12% increase of relative risk of OHCA for each 10 μg/m^3^ increase of PM 2.5 was found. A study on patients with implanted cardioverter-defibrillators suggests that PM 2.5 is associated with a higher risk of ventricular arrhythmias, both with an acute and an intermediate effect (4-, 5-, and 21-day exposure)^13^. Our results, showing that high air pollutants levels are associated with a higher risk of OHCA, confirm the above-mentioned results even considering a wider and unselected population without implanted cardioverter-defibrillators.

Moreover, a recent one-year study^24^ from a 7863 km^2^ area of Lombardy region conducted during 2019, highlighted a significant dose-response relationship between OHCA incidence and air pollutants (PM 2.5, PM 10, CO, NO_2_ and C_6_H_6_). More specifically, dividing the study period into days with high or low incidence of OHCA, according to the median value of the daily incidence (0.3 cases/100.000 inhabitants), authors found that the concentrations of most pollutants were significantly higher in the days with a greater number of OHCAs as compared to those with a lower incidence.

Compared to the above-mentioned results and previous literature, our study shows some novelty features. First, data were collected prospectively and derived from a highly polluted but relatively small and homogenous urban area. In fact, monitoring stations data and OHCAs of residents in Piacenza City resulted from an area of 118 km^2^, with the same altitude, temperature, urbanization, population density and meteorological conditions; that could result in a greater sensitivity, thereby removing possible confounders related to wider or non-homogeneous area as well as to events occurred in non residents. Second, due to the small area considered in our study, it was possible not only to compare the number of OHCA per day with air pollutants concentration but also to compare days without OHCA and days with OHCA. Third, our study covers a long period (7 years), investigating five different air pollutants and either separately short-mid- and long-term exposure. Finally, ECG tracings and rhythm of all patients who underwent cardiac resuscitation were collected and separately presented.

Regarding long-term exposure to air pollution, fine particulate air pollution has been associated with ischaemic heart disease and stroke mortality; each increase of 10 μg/m^3^ PM 2.5 was associated, in fully adjusted models, with a 16% increase in mortality from ischaemic heart disease and a 14% increase in mortality from stroke^25^. Moreover, Montone et al. recently provided ^6^ novel insights into the missing link between air pollution and increased risk of coronary events. In particular, the authors found that long-term exposure (2 years) to higher PM concentrations is associated with the presence of vulnerable plaque features, plaque rupture and plaque inflammatory activation as a mechanism of coronary instability. Although the results by Montone et al. ^6^ shed new light on the mechanisms underlying the relationship between pollutants and plaque instability, our results, suggest for the first time that shorter exposure to air pollutants may precipitate life threatening arrhythmic events.

Finally, although the present study cannot provide mechanistic data capable of justifying the relationship between pollution and cardiac arrest risk, some pathophysiological considerations should be suggested. Since sudden cardiac death represents the acute fatal consequence of the association between arrhythmogenic cardiac substrate (e.g. ischemic heart disease, cardiomyopathy, channelopathy, hypertrophy) and pro-arrhythmic trigger (eg dysionia, dysautonomia, acute plaque rupture, acute decompensated heart failure, acute free radicals damage), the mechanisms linking pollution and sudden cardiac death can be multiple. In fact, recent evidences have associated acute or chronic exposure to environmental pollutants with many of the above-mentioned factors, such as dysautonomia^26^, endothelial dysfunction^27^, hypertension^28^, inflammation^29^, hypercoagulability^30^, coronary vasomotor disorders^31^, plaque instability^6^ and ultimately fatal arrhythmias^13^.

Our study has some limitations. Firstly, we lack of anamnestic data before OHCA. Secondly, we cannot exclude that some cases of asystole were the primary cardiac arrest rhythm not necessarily preceded by ventricular tachycardia or ventricular fibrillation thus suggesting a non-ischemic cause of OHCA. Thirdly, individual exposure to air pollution was assumed from the exposure in the area of the events like previous epidemiological studies although recent devices (air pollution trackers) have been recently proposed for measuring individual exposure. Finally, we cannot extrapolate from this data if PM acts mainly through plaque destabilization versus myocardial arrhythmias triggering. The role of PM on susceptibility to fatal ventricular arrhythmias should be assessed in future studies.

In conclusion, short-term and mid-term exposure to PM 2.5 and PM 10 is independently associated with a higher risk of OHCA due to asystole or shockable rhythm.

## Data Availability

The data that support the findings of this study are available from the corresponding author, upon reasonable request.

---

**Figure S1.**
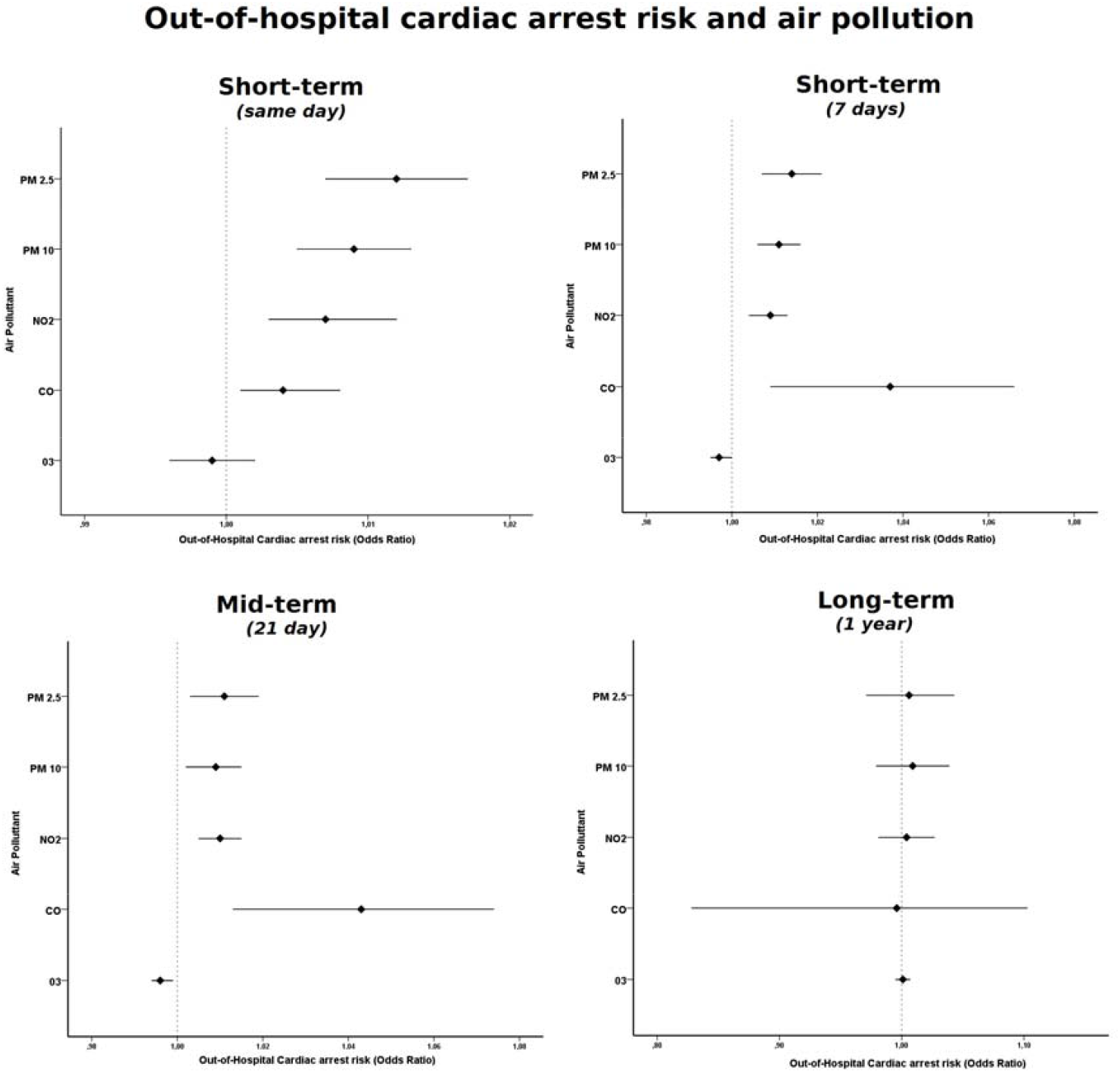
Short-term (same day), mid-term (7 days and 21 days), long-term (365 days) of air pollution exposure and out-of-hospital cardiac arrest (OHCA) risk.

**table S1.** Air pollution and OHCA risk according to different periods of exposure

|  | **Short term**<br>(7 days) |  |  | **Mid-term**<br>(21 days) |  |  | **Long-term**<br>(365 days) |  |  |
| --- | --- | --- | --- | --- | --- | --- | --- | --- | --- |
|  | **OR** | **IC (95%)** | **p** | **OR** | **IC (95%)** | **p** | **OR** | **IC (95%)** | **p** |
| **PM 2.5** | 1.014 | 1.007-0.021 | <0.0001 | 1.011 | 1.003-1.019 | 0.009 | 1.006 | 0.971-1.043 | 0.734 |
| **PM 10** | 1.011 | 1.006-1.016 | <0.0001 | 1.009 | 1.002-1.015 | 0.009 | 1.009 | 0.979-1.039 | 0.569 |
| **CO** | 1.037 | 1.009-1.066 | 0.001 | 1.043 | 1.013-1.074 | 0.004 | 0.956 | 0.828-1.103 | 0.537 |
| **NO<sub>2</sub>** | 1.009 | 1.004-1.013 | <0.0001 | 1.010 | 1.005-1.015 | 0.001 | 1.004 | 0.981-1.027 | 0.754 |
| **O<sub>3</sub>** | 0.997 | 0.995-1.000 | 0.059 | 0.986 | 0.994-0.999 | 0.011 | 1.001 | 0.955-1.007 | 0.073 |

Legend. OHCA, out-of-hospital cardiac arrest; PM, particulate matter; O<sub>3</sub>, ozone; CO, carbon monoxide; NO<sub>2</sub>, nitrogen dioxide; OR, Odds Ratio, CI, confidence interval.

**Table S2.** Air pollution and OHCA risk according to age and gender

|  | Odds Ratio | IC | P |
| --- | --- | --- | --- |
| **PM 2.5** | 1.012 | 1.007-1.017 | <0,0001 |
| Age < 65 years | 1,008 | 1,015-1,002 | 0,009 |
| Age > 65 years | 1,017 | 1,024-1,010 | <0,0001 |
| Male gender | 1,016 | 1,022-1,009 | <0,0001 |
| Female gender | 1,008 | 1,015-1,001 | 0,029 |
| **PM 10** | 1.009 | 1.005-1.013 | <0,001 |
| Age < 65 years | 1,006 | 1,011-1,001 | 0,018 |
| Age > 65 years | 1,013 | 1,019-1,008 | <0,0001 |
| Male gender | 1,012 | 1,017-1,007 | 0,0001 |
| Female gender | 1,005 | 1,011-1,000 | 0,065 |
| **CO** | 1.456 | 1.124-1.886 | <0.001 |
| Age < 65 years | 1,028 | 1,061-0,995 | 0,093 |
| Age > 65 years | 1,052 | 1,091-1,015 | 0,006 |
| Male gender | 1,040 | 1,074-1,007 | 0,017 |
| Female gender | 1,035 | 1,072-0,999 | 0,058 |
| **NO<sub>2</sub>** | 1,007 | 1.003-1.0012 | <0,001 |
| Age < 65 years | 1,006 | 1,011-1,001 | 0,02 |
| Age > 65 years | 1,010 | 1,015-1,004 | <0.001 |
| Male gender | 1,008 | 1,014-1,003 | <0.001 |
| Female gender | 1,006 | 1,012-1,001 | 0,028 |
| **O<sub>3</sub>** | 0,999 | 0.996-1.002 | 0.577 |
| Age < 65 years | 0,998 | 1,001-0,995 | 0,296 |
| Age > 65 years | 0,998 | 1,001-0,994 | 0,188 |
| Male gender | 0,998 | 1,001-0,994 | 0,141 |
| Female gender | 0,999 | 1,002-0,995 | 0,418 |

Legend. OHCA, out-of-hospital cardiac arrest; PM, particulate matter; O<sub>3</sub>, ozone; CO, carbon monoxide; NO<sub>2</sub>, nitrogen dioxide; OR, Odds Ratio, CI, confidence interval.

